# A National Survey Exploring Dementia Care Provision in Social Care in England

**DOI:** 10.64898/2026.11.09.26367668

**Authors:** Ayoni Darby, Conor Durkin, Hannah Chapman, Oliver Kelsey, Charlotte Kenten, Sube Banerjee, Yvonne Birks, Claudia Cooper

## Abstract

**Context:** Dementia is associated with significant use of social care, yet we know little about how providers respond to increasing demand and policy ambitions to deliver more care in community settings.

**Objective:** To examine dementia care provision, workforce training, service integration, and perceived challenges among care homes and home care agencies in England.

**Methods:** We conducted a mixed methods co-designed survey of care home and home care agency managers across five geographically diverse English regions, exploring: training, service delivery, relationships with other services, and future priorities. We analysed data descriptively and free-text responses using qualitative content analysis.

**Findings:** We received valid responses from 42 home care agencies and 47 care home managers, who estimated around one third of home care clients, and most care home residents, had significant cognitive impairment. Most care homes held regular multidisciplinary meetings (29/40, 72.5%). Home care managers described positive collaborations with health and other social care services, but also difficulties accessing support and concerns about undervaluation of care workforce skills within multidisciplinary care. Across both settings, frequently reported challenges included: increasing care complexity, behaviours that challenge, recruitment, retention and financial constraints. In home care settings, difficulties working in partnership with families were cited as reasons for care breakdown. Most organisations provided dementia-specific training, identifying communication, behavioural support, de-escalation, safeguarding, and working with families as unmet training needs.

**Limitations:** Findings are probably limited by response and desirability bias; we did not triangulate findings with objective data.

**Implications:** As policy shifts care to community settings, investment in social care workforce development, specialist support and integrated pathways are essential for sustainable high quality dementia care. Increased funding, improved training, stronger career pathways, and greater recognition of care workers were identified as priorities for improving dementia care.

## Introduction

The number of people with dementia is rising worldwide; it is one of the most pressing global health and social care concerns. In England, 498,926 people were diagnosed with dementia in 2025, and it is the leading cause of death (Nuffield Trust, 2026). Dementia profoundly affects individuals, families, and wider society, with UK annual costs expected to reach £90 billion by 2040; most costs are borne by families and social care (Alzheimer’s Society, 2024). Almost 400,000 people live in English care homes, and a further half a million receive local authority funded home care (GOV.UK, 2026). Approximately 60% of people receiving in-home social care in England (United Kingdom Homecare Association, 2015), and three-quarters of care home residents are estimated to live with dementia (Livingston et al., 2017), supported by 900,000 social care workers (Hopkins, 2025).

Dementia care should be personalised, enabling, inclusive and collaborative (Dalgarno et al., 2021; Lord et al., 2020), but a Care Quality Commission (CQC) commissioned audit, involving 53 social care providers (including home care agencies and care homes), reported that current dementia training in these settings insufficiently equips the workforce to deliver high-quality person-centred care (Smith et al., 2026). The report reflected a key workforce challenge: care workers are low paid, often undervalued and expected to provide skilled care to people with typically just 1-2 hours of dementia specific training (Brown et al., 2022; Leverton et al., 2021a, 2021b; Smith et al., 2026). An accompanying survey of 184 social care workers, found, when benchmarking provision against the Dementia Training Standards Framework (Skills for Health, 2018) that most training delivered was at Tier 1 (awareness level), and there were critical gaps at Tier 2, for example training around equality, diversity and inclusion, families as partners in care, and end of life care (Smith et al., 2026).

The *Fit for the future:10 Year Health Plan for England* (2025) asks the health system to deliver three “big shifts”: from sickness to prevention, hospital to community and analogue to digital. It acknowledges the importance of social care and promotes greater health and social care integration. However, broader social care funding and structural reform remain largely outside its scope despite the acknowledgement that social care will be critical to this shift, especially among people with dementia who often have complex needs (Alzheimer’s Society, 2024).

Developing an appropriately skilled social care workforce is therefore a policy priority (NHS England, 2023). The UK government’s long-term vision is for a National Care Service, to deliver consistency of care (UK Parliament, 2024). The Casey Commission is investigating the adult social care landscape and will make medium-term recommendations (expected 2026) to deliver *“funding flows and accountability mechanisms to improve quality and productivity”* (Casey Commission, 2025). Plans are likely to focus on appropriate funding and fair pay, and training and support for care workers to improve staff retention, wellbeing and drive-up care quality.

In relation to dementia specifically, a recent review found evidence for the effectiveness of dementia training in care homes in reducing agitation and improving quality of life; with some evidence for training effectiveness in home care. Approaches that engage staff “champions” to integrate practice-based learning were key, highlighting the critical role of the provider network in delivering better dementia social care (Delray et al., 2024). Investment to implement this evidence could deliver savings across health and social care from reductions in acute admissions and GP visits.

National surveys exploring how care home staff manage behaviours that challenge (Mallon, Krska & Gammie, 2019), sensory loss (Andrusjak, Barbosa & Mountain, 2021) and report neglect and abuse (Cooper et al., 2018) among people with dementia, highlight the diverse challenges providers face. To understand the challenges of the contexts in which social care workers support clients with dementia, we developed two co-designed survey instruments for care home and home care service managers and senior staff.

Within a broader national research project exploring dementia provision within differing health and social care organisations, we surveyed domiciliary and residential service managers in diverse areas of England (Kelsey et al., 2025; Muralidhar et al., 2025). We aimed to examine dementia care provision, workforce training, service integration, and perceived challenges. Our objectives were to understand how social care organisations:

1. Plan and deliver staff training and provide inclusive, high-quality care to service users.
2. Enable, or are unable, to deliver care integrated with local health service provision.
3. Perceive key challenges, as well as factors that generate pride and concern.

### Methodology

This cross-sectional national provider survey was conducted by the NIHR Dementia and Neurodegeneration Policy Research Unit at Queen Mary, University of London (DeNPRU-QM), commissioned by the Department of Health and Social Care (DHSC) and approved by the National Research Ethics service and Health Research Authority on 14.05.2024 (24/IEC08/0008).

#### Survey development and co-design

We developed the surveys (one addressing each context) using a co-design approach with people with lived experience of dementia, and service managers or staff deputising for them from diverse regions of England.

The survey was co-designed around the study objectives outlined above, with input from stakeholder groups. The DeNPRU-QM Patient and Public Involvement and Experience (PPIE) group (seven PPIE members living with or caring for a person with dementia) met with CC and CK in an online focus group, to share ideas about survey development and guide initial survey drafting by the research team. We then held an in-person workshop with four home care agency managers, a senior manager and a home care worker on 30.04.24; and an online workshop with two care home managers, and care home worker and a senior leader in a care home franchise; four nurses, a junior doctor, and psychiatrist, on 29.06.24. Before these meetings, participants were sent an evidence pack to provide context and the drafted survey. During meetings, members proposed and discussed survey items in line with the evidence and aims, providing feedback using a structure, process and outcome framework. After the co-design groups, the drafted surveys (for managers of care homes and home care agencies respectively) were revised using participants comments, who were invited to review revised versions before the final version was produced using Qualtrics (www.qualtrics.com).

The surveys asked about organisational structure (staff and registration) and staff retention; staff training, and processes for managing client care, and ensuring services are inclusive. They comprised descriptive, discrete choice closed and open questions with free text responses for further expansion (surveys are provided in supplementary material). For questions regarding behaviour that challenges, we drew on previous work with care workers (Wong et al., 2025).

#### Survey sample and data collection

We selected five geographically diverse Integrated Care Boards (ICBs) areas in England within which to conduct the survey, in London, Midlands, South East, and two in North East and Yorkshire regions. Selected urban, rural and coastal regions encompassed diverse ethnic and socioeconomic populations. We used publicly available CQC data to identify eligible home care agencies and care homes.

Contact was through the National Institute of Health (NIHR) ENRICH network and then directly by AD and CD. Providers were sent a cover letter, participant information sheet (PIS) and link to complete the survey independently online via Qualtrics, or if they preferred on the phone with a researcher. Consent was completed electronically prior to survey completion. A £15 voucher was offered as renumeration and only one participant was included per organisation.

We invited one senior member of staff (usually the manager) with a deep knowledge of their service to complete the survey. Participants were currently employed by care homes or home care agencies within the five specified ICBs. Our intention *a priori* was to purposively select managers from 75 of these organisations to reflect diversity of organisation size, and to include franchises and standalone organisations.

#### Analysis

We exported survey data from Qualtrics to Microsoft Excel. Survey responses were included if they contained the participant demographics and a minimum of 5 completed survey questions, agreed as a requirement by the team to be inclusive but to exclude surveys that did not include sufficient responses to meaningfully capture the participant’s views.

We used standard descriptive statistics to summarise responses to fixed choice items; where items described a range, the midpoint in analyses was used (e.g. 5-10 became 7.5) and if greater or less than signs were written a minimum estimate was assigned (for example >20 became 21. We reported medians and interquartile ranges (IQR) of continuous data, as a skewed distribution was expected due to variances in service size and workforce characteristics using Excel and RStudio (Posit team, 2025).

We processed free text responses in Microsoft Excel, conducting qualitative content analysis. AD and CD coded all home care and care home responses, respectively. CC then independently coded 10% of responses for each question, using the coding list developed, discussing any discrepancies in codes and iteratively modifying the coding frame. We used frequency counts and salient quotes to summarise the prevailing themes amongst those that had written narrative comments.

## Results

Of the 236 services that accessed the questionnaire, 89 (38%) provided valid responses and were included in our analyses: those deemed invalid had not completed the required number of survey questions (see above). Almost all excluded surveys were blank and on review did not include usable responses to other questions that would have enhanced or changed our findings. 42/115 (37%) of home care agency responses received, and 47/121 (39%) of care home responses were valid with varying individual response rates for each question resulting in differential denominators.

### Provider and workforce characteristics (Table 1)

#### Home care agencies

Valid responses received were from: London (n=8), Midlands (n=9), Southeast (n=7), North East (n=8) and Northwest English regions (n=10). Most survey respondents were the agency manager/ director (n=36, 85.7%); half had been in the role at least five years (n=22, 52.4%), and four (9.5%) for less than a year. Around half of participating agencies were single site, and half were one branch of a multisite provider or franchise (which included a median of two sites, IQR): 1-4).

**Table 1:**
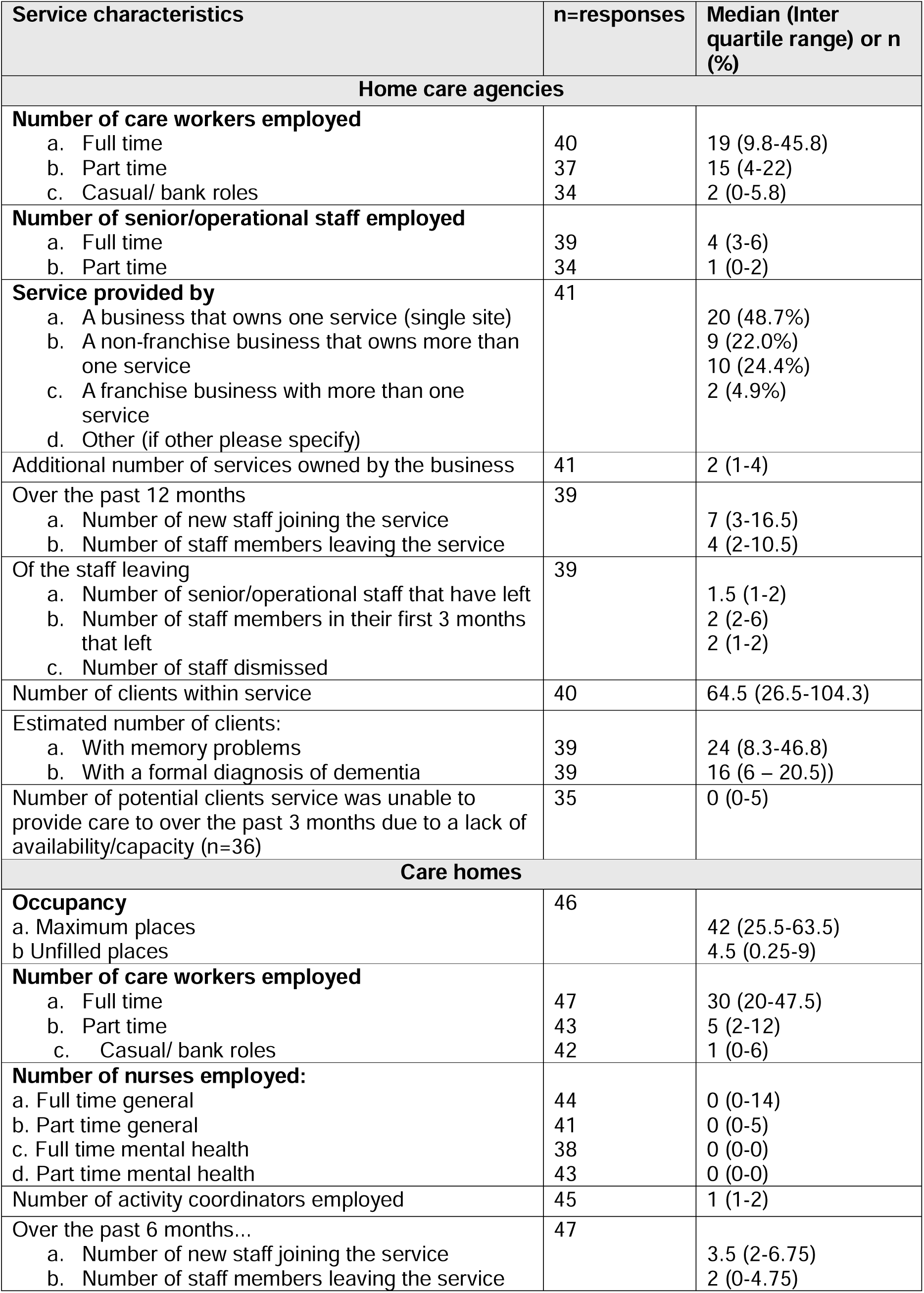
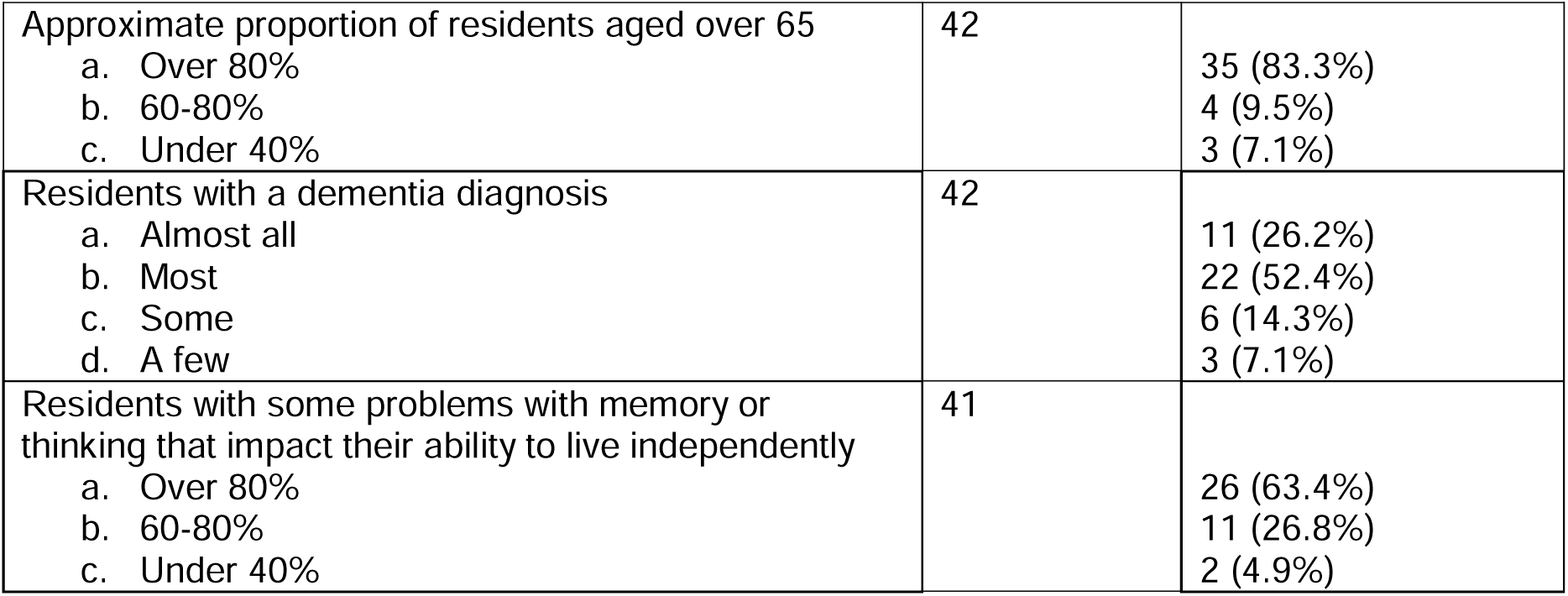
Characteristics of provider organisations represented by participants.

| Service characteristics | n=responses | Median (Inter quartile range) or n (%) |
|---|---|---|
| **Home care agencies** |  |  |
| **Number of care workers employed** |  |  |
| a. Full time | 40 | 19 (9.8-45.8) |
| b. Part time | 37 | 15 (4-22) |
| c. Casual/ bank roles | 34 | 2 (0-5.8) |
| **Number of senior/operational staff employed** |  |  |
| a. Full time | 39 | 4 (3-6) |
| b. Part time | 34 | 1 (0-2) |
| **Service provided by** | 41 |  |
| a. A business that owns one service (single site) |  | 20 (48.7%) |
| b. A non-franchise business that owns more than one service |  | 9 (22.0%) |
| c. A franchise business with more than one service |  | 10 (24.4%) |
| d. Other (if other please specify) |  | 2 (4.9%) |
| Additional number of services owned by the business | 41 | 2 (1-4) |
| Over the past 12 months | 39 |  |
| a. Number of new staff joining the service |  | 7 (3-16.5) |
| b. Number of staff members leaving the service |  | 4 (2-10.5) |
| Of the staff leaving | 39 |  |
| a. Number of senior/operational staff that have left |  | 1.5 (1-2) |
| b. Number of staff members in their first 3 months that left |  | 2 (2-6) |
| c. Number of staff dismissed |  | 2 (1-2) |
| Number of clients within service | 40 | 64.5 (26.5-104.3) |
| Estimated number of clients: |  |  |
| a. With memory problems | 39 | 24 (8.3-46.8) |
| b. With a formal diagnosis of dementia | 39 | 16 (6 – 20.5)) |
| Number of potential clients service was unable to provide care to over the past 3 months due to a lack of availability/capacity (n=36) | 35 | 0 (0-5) |
| **Care homes** |  |  |
| **Occupancy** | 46 |  |
| a. Maximum places |  | 42 (25.5-63.5) |
| b Unfilled places |  | 4.5 (0.25-9) |
| **Number of care workers employed** |  |  |
| a. Full time | 47 | 30 (20-47.5) |
| b. Part time | 43 | 5 (2-12) |
| c. Casual/ bank roles | 42 | 1 (0-6) |
| **Number of nurses employed:** |  |  |
| a. Full time general | 44 | 0 (0-14) |
| b. Part time general | 41 | 0 (0-5) |
| c. Full time mental health | 38 | 0 (0-0) |
| d. Part time mental health | 43 | 0 (0-0) |
| Number of activity coordinators employed | 45 | 1 (1-2) |
| Over the past 6 months... | 47 |  |
| a. Number of new staff joining the service |  | 3.5 (2-6.75) |
| b. Number of staff members leaving the service |  | 2 (0-4.75) |
| Approximate proportion of residents aged over 65 | 42 |  |
| a. Over 80% |  | 35 (83.3%) |
| b. 60-80% |  | 4 (9.5%) |
| c. Under 40% |  | 3 (7.1%) |
| Residents with a dementia diagnosis | 42 |  |
| a. Almost all |  | 11 (26.2%) |
| b. Most |  | 22 (52.4%) |
| c. Some |  | 6 (14.3%) |
| d. A few |  | 3 (7.1%) |
| Residents with some problems with memory or thinking that impact their ability to live independently | 41 |  |
| a. Over 80% |  | 26 (63.4%) |
| b. 60-80% |  | 11 (26.8%) |
| c. Under 40% |  | 2 (4.9%) |

Agencies employed a median of 19 full time (9.8-45.8 IQR), 15 part time (4-22 IQR) and 2 bank (0-5.8) members of staff (see Table 1). In the past 12 months, a median of four staff left each domiciliary provider (IQR 2 – 10.5); annual staff turnover in the agencies reporting data was 10.4%.

#### Care homes

We received 47 valid survey responses from: Northwest England (n = 10), North East England (n = 12), Midlands (n = 9), the South East (n = 8), and North East London (n =8). Care homes provided residential (n = 28), nursing (n = 11), or residential and nursing care (n = 8). Most were privately owned (44, 93.6%); the remainder (3, 6.3%) were “not for profit”. Around half were a standalone business (25/47, 53.2%), while the others were part of a chain, broadly reflecting the provider landscape in England (Blackwell, 2026).

Responders were mostly care home managers/directors (n = 37), while four were deputy managers. Other roles included a senior nurse, senior care assistant, human resource manager, administrator, team leader, home coordinator, and cognitive stimulation therapist. Around a fifth of participants had been in their role less than a year (n=10, 21.3%), 11 for 1-3 years (23.4%), 13 for 3-5 years (27.6%), and 13 for five years or more (27.6%).

A median of 40 (IQR: 30-64.5) staff were employed by care homes who responded. As shown in Table 1, most staff were fulltime carers. As expected, at least two nurses were employed in the 19 homes that provided nursing care. Nine of these homes employed at least one mental health nurse and two employed two; 18 employed a general nurse. In the past 6 months, a median of two staff (IQR: 0-4.75) had left their provider, equating to an approximate annual staff turnover of 10%.

### Reason for staff attrition and workforce concerns

Within home care services, reasons why staff left were grouped during content analysis and included: alternate job opportunities or further education (5/37, 13.5%), “*such as upskilling to nursing roles or pursuing opportunities within the NHS*”, travel and “*mileage costs*” (4/37, 10.8%), inflexible, unsociable hours (4/37, 10.8%), and low pay and cost of living crisis (3/37, 8.1%), with one manager of an urban, multi-site agency citing the cost of living crisis, clean air zone charges, progression to the NHS sector and general career changes. When asked to identify concerns around how their care home had operated in the past five years, staffing retention and a reliance on agency workers was most frequently raised (8/37, 21.6%) with one manager commenting:

*“Struggled a little bit for staff. Night staff might bicker, not turn up etc. Since they got to know the agency, they get given the same staff e.g. for 3 years. Relationship [is] a lot better now. The agency [agency] know the residents well. Previously they had a tough time e.g. 7pm ringing to say they can’t come for the night shift. More expensive to have agency during the night but more reliable” (registered manager)*

### Client/resident caseload and need

Home care agencies had a median of 64.5 clients (IQR 26.5 – 104.3). Around a third of clients (median 24, IQR 8.3-46.8) were known to have memory problems and a quarter (16; 6-20.5) had a dementia diagnosis. In comparison, most care homes reported that over four-fifths of residents were aged 65 or older and experienced some degree of memory or cognitive impairment that may affect their ability to live independently, and most or almost all had a dementia diagnosis (Table 1).

15/35 (42.9%) home care services indicated they were providing care to clients that were unable to speak English sufficiently to communicate their needs. In the free text comments, strategies to address this included matching clients to staff that spoke the same language (8/16), pictures or non-verbal methods such as common hand signals (6/16, 37.5%) and translations apps (5/16); only three used interpreters and two worked with family members or advocates. Care homes more frequently reported providing care to residents with limited English (n = 26/39). Six reported having access to a local authority funded interpreter, a further six accessed interpreters through privately funded arrangements, while most (n = 27/39) reported having no access to interpreter services. Approaches to managing language barriers were similar to those used by home care agencies, most frequently: communication techniques (n = 19/24), multilingual staff (n = 10/24) and translations tools/technology (n = 7/24).

Care homes provided a median of 10 (n = 40, IQR: 6.25-13.75) activities per week. Residents could take part in the activities even when the activity coordinator was not present in most care homes (35/41, 85.3%), while in the other six homes, this was dependent on the activity. The most cited activities included physical activity/exercise (n = 22/40), music-based/entertainment (n = 21/40), games/cognitive (n = 21/40), and creative/arts and crafts (n = 20/40).

### Reasons care could not be provided or sustained (Table 2)

#### Home care

Table 2 presents home care agency responses when asked about situations they were unable to meet the needs of clients with dementia, required additional support, or when care provision broke down. Common themes related to higher care needs, funding and working with family carers.

**Table 2:** Reasons home care clients with dementia could not be supported to live at home, and times when care broke down and what would have helped: content analysis.

| Theme | Participants supporting | Example of quote coded in theme |
|---|---|---|
| **Reasons clients could not be supported<sup>1</sup> (N=31)** |  |  |
| Higher care needs | 18/31 (58.1%) | “because of their safety and requiring 24 hours of support”<br>“client did go missing on one occasion” |
| No/ not applicable | 10/31 (32.3%) |  |
| Behaviours that challenge | 7/31 (22.6%) | “so aggressive to husband that she has attacked him and he has fallen and broken his hip”<br>“wife was physically abusive to her husband and caused harm” |
| Family carer related issues | 5/31 (16.1%) | “One was when the family had no appreciation of their loved one dementia and expected outcomes which were not viable”<br>“this person was eventually admitted to care home as the family were unable to provide the increased support necessary in-between our calls. |
| Restrictions on care packages/ costs | 4/31 (12.9%) | “restrictions places on care packages by local authority”<br>“2 carers 4 times a day however his social visits were removed with funding”<br>“financial consideration as to whether it is more cost effective to transfer to a care home than increase the time spent by carers in the community” |
| **Reasons for why care has broken down<sup>2</sup> (n=35)** |  |  |
| None/ not applicable | 9/35 (25.7%) | I don't believe there was any other help that would of changed the outcome” |
| Unable to establish collaborative care partnerships with families/ wider support networks | 7/35 (20%) | “gentleman who is living at home with his wife. It has become apparent that she also has dementia”<br>“most challenges we face regarding support[ing] a client with Dementia is the opinions of family members and their wishes”<br>“Family members removing the choice for people with dementia, and dismissing their right to choice”<br>“sister wouldn’t engage”<br>“families feel they automatically need residential care” |
| Higher care needs | 5/35 (14.3%) | “he has got lost in the past. Safeguarding [has] been raised and currently active” |
| Finances | 5/35 (14.3%) | “clients family moved to a cheaper provider”<br>“reluctance to increase the package of care due to the cost” |
| Client carer relationship breakdown | 2/35 (5.7%) | “accusations made about staff”<br>“abusive towards workers and family” |
| **Suggestions for what could have prevented care breakdown<sup>2</sup> (n =35)** |  |  |
| More funding/local authority support | 7/35 (17.1%) | “greater funding/ financial support for people requiring help with regulated activities such as personal care. Sadly many self-funders are limited by what care they can sustainably afford”<br>“Allowing longer visit times so care was not rushed”<br>“care needs assessment at the time of admission coupled with risk assessments is of prime importance… instances were the commissioned packed of care is not person centred” |
| More support for families/ family involvement | 5/35 (14.3%) | “Family understanding what the diagnosis means and how best to cope with it from their perspective and from the carers. Listening to what we are explaining to them and understanding that their mum or dad is there, just being masked by this behaviour”<br>“earlier conversations with families about the likely progression” |
| More training/ experienced staff | 5/35 (14.3%) | “difficult to recruit experienced carers”<br>“proper dementia specific training for staff on communication and behaviour support” |
| More specialist support/ step up options/ signposting | 5/35 (14.3%) | “more rapid access to specialist dementia support, including night-time provision and crisis response services…clearer escalation pathways”<br>“greater availability of intermediate or step-up care options”<br>“more services and organisations like the Alzheimer’s Society” |

<sup>1</sup>Survey question 11: “Have there been clients with dementia who you have been unable to support living at home longer or where additional help was needed? If so, please tell us about this:”
<sup>2</sup>Survey question 12: “Tell us about times where care for people with dementia has broken down/ not worked out and what would have made a difference?”

Higher care needs with clients requiring a care home or hospital bed were referenced by 18 (n=31) respondents as a typical reason why clients could not be supported by domiciliary care. One home care agency manager from a franchise in the Midlands region wrote:

*“a gentleman was wandering at night and went missing, creating a significant and ongoing safeguarding risk. While options such as the Herbert Protocol and door alarm systems were carefully considered, these measures were ultimately insufficient to manage the level of risk safely.”*

Seven respondents (n=18) gave behaviours that challenge, including physical violence, as a reason for individuals could not be supported. Participants described relationship breakdown between clients and workers, with clients being “*abusive towards workers*” and making “*accusations*” about staff. Aggressive behaviours were also noted in responses to closed survey questions (with two thirds (19/28, 67.9%) of respondents reporting experiencing some from of abuse from a client (in 12 cases physical abuse) and 50% (10/20) from a family carer in the last three months.

Failure to establish collaborative partnerships with family carers or wider support networks were described as a challenge to care provision:

*“the family had no appreciation of their loved one’s dementia and expected outcomes which were not viable.”*

“*most challenges we face regarding support[ing] a client with Dementia is the opinions of family members and their wishes*”

While increasing care needs might be theoretically met by increased care provision, the challenge of financial and capacity ceilings was highlighted by several providers:

“*this person was eventually admitted to a care home as the family were unable to provide the increased support necessary in-between our calls. Often there is a financial consideration as to whether it is more cost effective to transfer to a care home than increase the time spent by carers in the community*”

*“Sister had the lady’s money but wouldn’t pay – social didn’t want to pay. We had to say we are pulling out until it’s done. It was upsetting for all”*

When asked what could have prevented care breakdown, respondents cited: more funding or local authority support (7/35), greater support for families or more family involvement in care (5/35), more training or experienced staff (5/35) and more specialist support/ step up options (5/35):

*“ … What would have made a difference was: Ensuring continuity of carers so he could build trust and familiarity. A more person-centred care plan. Proper dementia-specific training for staff on communication and behaviour support. Involving the family earlier to gather insight … Allowing longer visit times so care was not rushed.”*

#### Care homes

The most common reasons provided for care homes not accepting clients in the last six months were behaviours that challenge (18/39); a care home manager noted:

“*Unsuitable placements, referred by mental health teams, such as highly challenging, violent, and aggressive…”*

Additional reasons were: the provider deciding they could not meet the person’s complex needs (n = 13/39), or specific nursing needs (n = 9/39), for example for a percutaneous endoscopic gastrostomy (PEG), or oxygen therapy; not meeting entry criteria (n = 5/39) and “lack of funding” (n = 4/39). Other responses were “*concern[s] about compatibility with the existing resident group” (*2/39) or a lack of availability of ground floor rooms for those with poor mobility (3/39).

### Dementia training

#### Home care

All home care agencies responding provided some dementia specific training; 22/40 responses (55%) highlighted only training in house, 8/40 (20%) through external suppliers only, and 10/40 (25%) offered both. Twenty-four participants contributing useable data, provided a median of 14.5 hours (IQR 5 – 20.8) of dementia specific training. Most agencies delivered at least some in person training (84%, n=33), and e-learning (69.2%, n=27); six (15.4%) were entirely reliant on online/E-learning.

Eight (n=32) respondents were unable to identify any shortcomings in available training. Gaps that were highlighted in free text responses included practical, case-based, interactive training and experiential learning formats; *“Virtual reality simulations or the dementia bus which would help staff better understand how disorientating and uncomfortable living with dementia can be.”*

Decision making and communicating, de-escalation/ safeguarding and risk reduction were topics felt to be missing from existing provision:

*“Scenario-based training on de-escalation, communication in advanced-stage dementia, and supporting families through crisis periods would strengthen staff confidence further. Similarly, more detailed training around capacity assessments, best interest decision-making and safeguarding considerations specific to dementia would support safe practice.”*

#### Care homes

Most care homes reported that 90% or more staff receive dementia specific training (37/41, 90.2%). The median number of hours of in-house training provided to staff was 2 hours (n = 14, 2-18), plus a median of 4.5 (n = 11, 3-12.5) hours by an external supplier. Training was most often supplied with a mixture of in-person and e-learning sessions (23/41, 56.1%) followed by e-learning only (12/41, 29.2%) and in-person only (n = 6, 14.6%), most respondents considered current dementia training sufficient (n = 20/31, 64.5%). Similarly to home care respondents, gaps identified were in the managing and understanding of behaviours that challenge was most frequently highlighted as a gap in dementia training (n = 5/31, 16.1%):

*“Understanding reasons behind behaviours, triggers, causes and management”*

Two respondents (6.4%) wanted more interactive training:

*“I have been to the local hospital where they did training and you could see and act out what it feels like having dementia [by] interacting with the tutors, who were very knowledgeable”*

Most care homes offered support to staff when residents exhibited behaviours that challenge through various combinations of multidisciplinary discussion groups, additional training specific to behaviours that challenge and employing specific models of care. The most cited combination (n = 12/41, 29.2%) was that of regular multidisciplinary discussions and additional training specific to behaviours that challenge.

### Relationships with other services and multidisciplinary working

#### Home care

When asked about relationships with a range of other services, relationships with local authority adult social care services were most frequently described in positive terms within qualitative responses (24/36, 66.7%), followed by primary care (21/36, 58.3%), and dementia-related mental health services (15/36, 41.7%). A small number of respondents highlighted a shared commitment to their clients (5/42, 11.9%), and positive approaches to joint working as reasons for relationships working successfully:

“*In many cases, there is a shared commitment to maintaining individuals safely within the community and avoiding unnecessary escalation to acute setting”*.

*“Named social workers and regular contact points greatly improve continuity and coordination.”*

Where problematic relationships were reported, poor accessibility to, or inadequate communication with other formal services was noted:

“*Poor - difficult to get hold of allocated workers, or the case has been handed other to another worker, and we haven’t been informed and don’t know who to contact.”*

*“they do not listen”*

The depth of knowledge statutory social services had of dementia care was also questioned:

*“level of understanding to do dementia is limited”*.

Some respondents felt that the wealth of knowledge that home care workers held about clients was unappreciated, and that other services failed to appreciate the dynamic nature of support provided by social care providers:

*“Primary care providers also don’t see the importance of care providers, and understand the knowledge that we hold surrounding our clients.”*

*“Some OTs (occupational therapists) state that they have already been out and assessed. They seem to not realise that people’s mobility can change. We sometimes have to beg for moving and handling plans and they don’t understand why we need them*”

When asked which services home care agencies would like to be able to refer clients directly, 7/20 (35%) thought dementia specific services would be beneficial, 4/20 (20%) mental health teams, 4/20 20%) mentioned allied health professionals and 3/20 (15%) highlighted the falls and frailty teams.

#### Care homes

Across 40 responses, 31(77.5%) care homes indicated that one GP cared for all residents, while in nine homes residents had individual arrangements. GP involvement was predominantly reported as regular rather than happening on request (as required) (regular: 27/37, 72.9% vs as required: 10/37, 27%). Where specified, weekly visits were most common (13/26, 50%). Other professionals that visited care homes included speech and language therapists (ad hoc: 35/37, 94.5% vs regular 2/37, 5.4%), physiotherapists (ad hoc: 33/37, 89.1% vs regular 4/37, 10.8%), occupational therapist (ad hoc; 34/36, 94.4% vs regular: 2/36, 5.5%), psychiatrists (ad hoc: 32/33, 96% vs regular 1/33, 4%), and mental health nurses (ad hoc: 26/33, 78.7% vs regular 7/33, (21.2%).

Most care homes in our sample reported holding regular multidisciplinary meetings (29/40, 72.5%). Multidisciplinary meeting attendance mostly included combinations of GPs, nurses, and care home staff (12/30, 40%). GP involvement (either alone or in combination) was present in most responses, and nurses were also commonly represented. Regular review of psychiatric medication was commonly reported 39/40 (97.5%,), most frequently by a GP (26/34, 76.4%), sometimes including local mental health services (21/34, 61.7%), family members or next of kin (17/34, 50%), care home staff and management (14/34, 41.1%) and the resident (9/34, 26.4%) *“…if they had capacity*”.

### Improvements and concerns in the last five years

#### Care homes

Care homes were asked how their organisations had been working over the past five years. The most frequently reported improvements over the last 5 years related to staffing (n = 12/37, 32.4%) particularly in recruitment and retention of overseas staff:

*“Overseas recruitment and integrating 35 overseas staff into the home and local community. This ensured we had sufficient staff to provide high-quality care without using agency staff” (registered manager)*

Care homes were also proud of the improvements in person-centred care (n = 8/37, 21.6% :

*“…we have moved away from 1 [one-size] fits all to individualise our care plans to be person centred which reflects each residents needs and preferences…”*

The adoption of digital systems and technology (n = 8/37, 21.6%) has “…*made a massive difference*”, leaving staff more time “*to be with the residents instead of sitting writing notes*”. Improvements in training and clinical practice (n = 8/37, 21.6%) were equally cited as areas care homes were proud in the last five years. This was followed by changes in culture and staff wellbeing (n = 6/37, 16.7%) that has improved the “*…overall wellbeing of the residents and staff*”, upgrades or maintenance of accreditation and reputation (n = 6/37, 16.7%) from the Care Quality Commission (CQC) and improvements in physical environment (n = 6/37, 16.7%). Some care homes were proud of their relationships and communication (n = 5/37, 13.5%) with “…local professionals” and “…residents and families” with one citing that they particularly valued “community involvement and fundraising events…”.

While most respondents reported no concerns around how their care home has operated in the last five years (17/37, 45.9%), the most frequently cited issues related to staffing (8/37, 21.6%) and agency use. The pressure from limited funding (5/37, 13.5%) concerned some care homes:

*“Mainly financial pressures due to a lack of funding and ever-increasing cost of living. We are determined not to lower standards, but this is proving harder than ever”*.

External healthcare support issues were highlighted in two responses:

*“since the pandemic I find it very difficult for a GP to visit when needed and we do struggle with support from other professionals at times and can get passed from pillar to post. It is very frustrating when we are trying to get the support for our residents and feel that each door we approach is closed in our faces.”*

### Anticipated changes and priorities for reform

#### Funding concerns

Funding concerns predominated in responses in care home and home care respondents. When asked about proposed changes to how services operate that participants found concerning, funding and budget cuts were most frequently cited by home care agencies (8/29, 27.6%) with one participant stating: “*A lack of sustainable funding means local authorities cannot pay a fair hourly rate for care. This has driven a focus on the self-funded market, but cost-of-living pressures are reducing affordability. At the same time, increases in the National Living Wage and National Insurance have significantly raised costs, squeezing margins and impacting sustainability.”*

Another commented “*Council are looking at reverting all AO2 (assistance of 2) visits down to AO1 (assistance of 1) …This is a massive safety concern and we have had several service users be put back down to AO1 and within a week they are deemed not safe with transfers so then back up to AO2 - this has been very disruptive for both staff and service users.”*

Ten participants (of 29 responding) stressed that anticipated improvements in the home care sector were funding dependent :

*“Our LA (local authority) is merging with a wider LA which will be more beneficial as they offer a “Dementia” rate fee. This will enable us to provide extra staffing to make the environment more safe.”*

Anticipated changes in the care home sector also mostly related financial pressures (7/32) especially in association of those in receipt of statutory funding:

*“reduction in funding by local authority for residents*”

#### Changes to care provision

When asked about anticipated changes services expect within the next five years, most respondents predicted greater collaboration with other services (16/30, 53.3%)

“*Increase integrated working…participate in joint reviews…expand our role in supporting hospital discharge pathways and preventing avoidable admissions*” (Table 3).

**Table 3:** Changes anticipated to care providers over the next five years.

| Code | n | Example quotes |
|---|---|---|
| **To how home care agencies interact with other services (N=30)** |  |  |
| More integrated working | 16/30 (53.3%) | “better communication and joint working”<br>“Increase integrated working…participate in joint reviews…expand out role in supporting hospital discharge pathways and preventing avoidable admissions”<br>“more healthcare tasks in the community” |
| Digitisation improving services | 5/30 (16.7%) | “Utilise secure digital care systems to enhance communication and real-time information sharing”<br>“online connection with GP services to make accessing client medical records easier”<br>“remote monitoring” |
| Greater expectations with less funding | 5/30 (16.7%) | “Improve services but on a reduced budget”<br>“being able to afford to keep the day centre part of this business open” |
| More cases | 1/30 (3.3%) | “Working with more people with dementia” |
| Further negative digitisation | 1/30 (3.3%) | “further digitisation of health and social care… harder than ever to speak to someone over the phone” |
| Training needs | 1/30 (3.3%) | “expectation by healthcare bodies that they can train once and expect that the training will then be cascaded by those they trained, with no adequate competencies in place…area where stronger links need to be forged and realistic expectations” |
| **To how care is provided to service users (N=29)** |  |  |
| Funding dependent | 10/29 (34.5%) | “to have the finances to adequately train and support staff”<br>“Our LA is merging with a wider LA which will be more beneficial as they offer a "Dementia" rate fee. This will enable us to provide extra staffing to make the environment more safe” |
| Personalised care | 6/29 (20.7%) | “focus will remain on deliver safe, compassionate, dignified and person-centred care”<br>“development of strengths-based and trauma-informed approaches” |
| Increasingly digital | 6/29 (20.7%) | “More technology to manage care needs.”<br>“secure information-sharing platforms are likely to become more embedded within routine practice” |
| Increased training | 5/29 (17.2%) | “further in depth training for the team in terms of Dementia Care and Palliative” |
| More complex clients | 4/29 (13.8%) | “more complex needs within the community” |
| Continued care improvements/ hard working staff | 4/29 (13.8%) | “Carers work to a very high standard”<br>“remain caring about those that we look after” |
| Collaboration | 2/29 (6.9%) | “increase co-production by involving service users and families”<br>“communication and collaboration” |
| Less person centred | 1/29 (3.4%) | “will become task oriented not based on the individual” |
| **Changes care homes anticipate over the next five years (n=32)** |  |  |
| Funding pressures | 7 /32 (21.9%) | “Reduction of funding by local authority for residents”<br>“Financial concerns, cost of living rising higher than fees paid by L/A [LA] and ICB” |
| Expansion/growth | 6 /32 (18.8%) | “We are increasing our occupancy from 58 - 84 beds”<br>“I would like to expand and diversify, I had been planning in opening [sic] a unit for residential Learning Disabled, where we can offer a form of supported living in a home environment” |
| Clients have changing, more complex care needs | 6/32 (18.8%) | “…we all can see a difference in [between] residential status, where people who should be [in] nursing [care] are classed as residential, meaning the senior care team have to work with individuals with multiple morbidities, this puts extra pressure on the team, [and] the nursing clients now have more serious conditions… clearly a growing trend of treat at home [care homes], and as such is placing more strain on the care home market”<br>“[hospital] Discharge team say the resident does not have nursing needs to save money and is not fair to staff or resident. Hopefully this will change” |
| Innovating/ personalising services | 6/32 (18.8%) | “recreate a post office in the home … this will give them purpose and [to] continue to buy items and pay for them”<br>“improving the service of CST and activities” |
| Staffing training and development | 5/32 (15.6%) | “Further investment in staff growth and development to ensure a high level of understanding around behaviours that challenge.” |
| Community and partnership relationships | 5/32 (15.6%) | “Mental health services is [are] poor now in time scale for visits, can anticipate this to get worse”<br>“continued [continue] improving links with community health services and admission avoidance strategies” |
| Technology/digitalisation | 4/32 (12.5%) | “More digitalisation, digital medication (e.g. online ordering), currently still have to manually write what has been given [and] what they need. Digital medical recording would be better, paperless” |
| Staffing challenges (recruitment/retention) | 4/32 (12.5%) | “I hope that my current staff continue to stay with us as we do not use agency” |
| Quality and rating improvement | 3/32 (9.4%) | “…improving our rating from good to outstanding”<br>“Outstanding CQC” |
| No changes | 2/32 (6.3%) | “No changes” |

<sup>1</sup>Home care survey question 17: “What changes do you anticipate in your service in the next 5 years: a. To how you work with other services? B. To how you provide care for your service users? C. Anything else?”
<sup>2</sup> Care home survey question 21: “What changes, if any, do you anticipate in your care home over the next 5 years?”

Six respondents anticipated more personalised support over the next 5 years, including the adoption of “*trauma -informed and culturally competent approaches”*, and a greater use of “*digital technology, remote monitoring…that enable earlier intervention, improved access, and better self-management of long term conditions”* .

Likewise, care homes foresee a shift towards more innovation and personalisation of services (6/32).

A shift towards more complex needs among clients was also raised by both home care and care home providers, as an anticipated result of the shift from hospital to community care. Six care home providers raised this, with one describing “*clearly a growing trend of treat at home [care homes], and as such is placing more strain on the care home market.*”

Expansion and growth of the care home into larger sites and more varied units, responding to this anticipated increase in need, also featured in six answers:

*“We are increasing our occupancy from 58 - 84 beds. We are developing the [a] site which will enable us to offer our services to the community”*.

#### Workforce support

When we asked what would make an important, positive difference to adult social care providers, twelve home care respondents referred to improved training, accreditation or strengthened career pathways and six to the need for carers to be more valued:

“*more investment into the care sector, more recognition of the value of care staff and national recognition of the training that is involved in becoming a carer.”*

Care home respondents described development of staff in terms of better training (5/32) as a priority, some noting challenges in staffing particularly around the recruitment and retention so staff (4/32). Additionally, the incorporation of technology and digitalisation (4/32, 12.5%) to improve “*…medical recording…*” and wider operations is expected to improve digital support for workers.

## Discussion

This national survey provides insight into how English care homes and home care agencies support people diagnosed with dementia. It highlights workforce, service delivery, and system-level challenges. Across both settings, respondents described strong commitment to providing person-centred care despite increasing complexity of client needs, workforce pressures, and financial constraints. Findings suggest that the ability of social care services to meet rising demand associated with dementia is influenced by the interaction between workforce capability, funding, service integration, and access to specialist support.

Home care managers estimated that around one-third of their clients had memory problems, fewer than the estimated 60% of home care clients believed to have some form of cognitive impairment nationally (United Kingdom Homecare Association, 2015). Due to our modest sample size this is difficult to interpret and may reflect differences between our respondents and the wider sector. It may also point to a genuine gap in information sharing or recognition: cognitive impairment may be present but not formally diagnosed or a diagnosis may exist yet not be shared with home care providers by health services. In contrast, most care home residents were reported to have dementia or significant cognitive impairment, perhaps indicative of a more impaired population, or of care homes having stronger relationships with healthcare professionals alongside more consistent identification, and more multidisciplinary input and information sharing relative to home care agencies.

Funding pressures emerged as a dominant theme. Respondents frequently described situations in which care packages were constrained by local authority funding arrangements or by the financial circumstances of self-funding individuals and families. These financial pressures influenced the quantity and quality of care that could be delivered, including visit duration, staffing levels, and access to enhanced support. Managers expressed concern that without sustainable investment, services may struggle to meet future demand while maintaining quality and continuity of care.

As policy continues to promote care closer to home and reduce reliance on hospital-based services, the social care workforce will be expected to support people with complex dementia-related needs. Our findings suggest that many providers are already operating at the limits of available resources and capacity, leaving limited capacity to realise the potential of care integration, for example around virtual wards as a way of avoiding distressing admissions for people with dementia (Wong, A. et al., 2025).

Behaviours that challenge were among the most frequently cited reasons why services were unable to support individuals safely at home or why care arrangements broke down. Such findings are consistent with previous research demonstrating that behavioural symptoms, including aggression are major drivers of caregiver burden, service use, and transitions into residential care (Costello, H., et al., 2019). This also points to an unmet training need particularly in home care provision.

Although most organisations reported providing dementia-specific training, the duration and format varied considerably, and managers identified important gaps in content. Respondents, especially from home care settings highlighted a need for more practical and interactive dementia training focused on communication, de-escalation, risk management, and supporting families during periods of crisis, suggesting that existing provision may not fully equip staff to respond to increasingly complex care needs. Previous work has highlighted limited alignment between training delivered in social care settings and the Dementia Training Standards Framework (Smith, S. J., et al., 2026). Experiential and practice-based learning approaches may be more effective than more didactic training programmes in supporting high-quality dementia care (Delray, S. et al., 2024). Peer learning groups are mandated for doctors as a condition of registration, and no health professional would be expected to provide client-facing care without training involving case-based, peer learning. Most home care workers have no such training (Cooper, C. et al., 2024; Kelleher, D. et al., 2024) and the provider model makes it hard to achieve, particularly in the home care workforce. Workforce development will be critical to realising the ambitions of integrated, community-based dementia care.

While many respondents described positive examples of collaborative working between independent provision and statutory social care providers and health services, others reported difficulties accessing health and social care professionals, delays in obtaining support, and limited recognition of the expertise held by care workers. Home care staff felt that their detailed knowledge of clients was sometimes undervalued during multidisciplinary decision-making processes. Effective integration requires not only structural reform but also recognition of the contribution social care providers make to assessment, monitoring, and ongoing support of people living with dementia.

Approximately half of home care agencies and two-thirds of care homes reported supporting people with limited English proficiency, yet access to professional interpreter services was uncommon. Providers relied on multilingual staff, family members, and translation technologies. Pairing clients with a small, familiar pool of staff may support relationship building and care continuity but this could create risk if staff members are unavailable or raise scheduling difficulties. Whilst these approaches offer practical solutions, they may not always offer consistency or the accuracy required for complex conversations about dementia care, decision-making, or safeguarding, potentially compounding disorientation and distress (Cooper, C. et al., 2017).

Managers reported challenges recruiting and retaining staff, frequently attributing departures to low pay, limited career progression opportunities, unsociable or inflexible hours and movement into better-remunerated health sector roles. Travel demands were recognised as negatively influencing retention as unlike NHS employers home care agencies are not required to reimburse workers for mileage and rising fuel prices alongside the introduction of clean air zones appear to have compounded this burden. Respondents also reported high levels of abuse from clients and family members highlighting workplace safety and emotional burden as important but under-recognised aspects of dementia care work (Kadri, A. et al., 2018). Together, these findings suggest that improving workforce retention is likely to require a combination of fair pay, enhanced training, professional recognition, career development opportunities, and improved employment conditions.

Participants anticipated that future dementia care provision would involve greater integration with health services, increased use of digital technologies, and more personalised models of care. These aspirations, which align with theoretical models of good quality dementia home care (Cooper, C et al. 2013), were accompanied by concerns about funding sustainability and workforce capacity. Successful implementation of policies aimed at shifting care from hospitals to community settings will therefore require parallel investment in the social care infrastructure needed to support these ambitions.

### Strengths and limitations

This study is among the largest contemporary surveys of dementia care provision across both home care and care home sectors in England and includes organisations from geographically diverse regions. Domiciliary care represents a substantial portion of national dementia support provisions but is less researched. The response rate was modest, raising the possibility of response bias, as well as desirability bias, as respondents may have been more engaged with dementia care than non-participants. Data were reported by managers and senior staff rather than frontline care workers or people with dementia and their families, so reflect organisational rather than direct experiences of care. Many questions relied on self-report and estimation which may be subject to recall or perception bias.

### Implications for policy and practice

The findings suggest that achieving national ambitions for community-based dementia care will require greater investment in the social care workforce, enhanced access to specialist dementia support, and stronger integration between health and social care services.

Workforce training, especially in the home care sector, should move toward practical, skills-based approaches focused on behavioural support, communication, and collaborative working with families. Policy initiatives aimed at improving pay, professional recognition, and career progression within social care may contribute to improved workforce retention and care quality.

## Conclusion

Social care organisations play a central role in supporting people with dementia across England. Despite examples of innovation, commitment, and collaborative practice, providers face persistent challenges related to funding, workforce capacity, training, and service integration. As policy continues to promote care closer to home, addressing these challenges will be essential to ensure that people with dementia and their families receive high-quality, person-centred support and that social care services remain sustainable in the face of growing demand.

## Data Availability

All data produced in the present work is available upon reasonable request to the authors.

## Acknowledgement

This research is funded through the NIHR Policy Research Unit: Dementia and Neurodegeneration Queen Mary University of London, reference NIHR206110. The views expressed are those of the author(s) and not necessarily those of the NIHR or the Department of Health and Social Care.

## Notes

### Competing Interest Statement

The authors have declared no competing interest.

### Author Declarations

Health Research Authority and Health and Care Research Wales approval of the project. IRAS project ID: 338398, Social Care Research Ethics Committee reference: 24/IEC08/0008. Sponsor Queen Mary University London

