## Supplementary Materials for "A National Survey Exploring Dementia Care Provision in Social Care in England"

**Supplementary Material**

**Supplementary Table 1: Changes anticipated to home care agencies within 5 years**

**(a) Home Care Agency Survey Questions**

| Topic | Questions |
| --- | --- |
| Service demographics | Service name  Service location  What area is service near to  Is this service a specialist dementia provider |
| Participant demographics | Name  Role  Duration in role |
| Service characteristics | 1.How many care workers are employed at your service   a. Full time b. Part time c. Casual/bank roles   1.b How many senior/operational staff?  a. Full time  b. Part time  2. Is the service provided by  a. A business that owns more than one service  b. A business that owns one service  c. A business that is part of a franchise and owns one service  d. A business that is part of a franchise and has more than one service  e. Other (if other please specify)  2b. How many more services does this business own?  3. How many clients do you have?  4. Please estimate how many clients:  a. Have memory problems  b. Have a formal diagnosis of dementia  c. Are 100% Self-Funded (as opposed to being funded in all or any part by adult social care or NHS Continuing Health Care)  5. How many potential clients was your service not able to provide care to over the last 3 months due to lack of availability/capacity?  6. Over the last 12 months  a. How many new staff have joined the service  b. How many members of staff have left the service  6b. Of those leaving, were any:   a. Senior/Operational staff   *How many senior/operational staff members?*   b. Staff that were in their first 3 months   *How many staff members who were in their first 3 months?*   c. Dismissed   *How many staff members were dismissed?*   d. None of the above   7. Are there any themes as to why staff leave the service |
| Training | 8.Thinking about dementia training you provide to home care workers  Who provides the training   1. In house 2. By external supplier   *Which external supplier provides the training?*  8b. How many hours of training are provided to staff?  8c. Is the training:   1. In person 2. Online 3. E-learning 4. Video call 5. Other   *If other please specify*  9. What additional training related to dementia do you think your staff need that isn’t currently available? |
| Work Conditions | 10a. How often in the last 3 months have you experienced any form of abuse by:  A client (0= never; 4=very often)  A family carer (0= never; 4=very often)  10b. How often in the last 3 months has any member of your service experienced physical abuse by:  A client (0= never; 4=very often)  A family carer (0= never; 4=very often)  11. Have there been clients with dementia who you have been unable to support living at home longer or where additional help was needed? If so, please tell us about this:  12. Tell us about times where care for people with dementia has broken down/not worked out and what would have made a difference? |
| Working Relationships | 13. How would you describe your working relationship with:  a. Primary Care  b. Local dementia mental health services?  c. Adult social services?  d. Are there service relationships you feel work particularly well or less well? Tell us more.  14a. Are there services that you cannot refer to directly where you feel it would be beneficial to be able to?   1. Yes 2. No   14b. If yes, which services?  15.Do you provide care to clients who do not speak English sufficiently to communicate their needs?  1. Yes  2. No  15b. If yes, how do you manage this? |
| Planning for the future | 16. Are there any proposed changes to how your service operate that concern you? Tell us about this?  17. What changes do you anticipate in your service in the next 5 years:  a. To how you work with other services?  b. To how you provide care for your service users?  c. Anything else?  18. Tell us one thing that would make an important and positive difference to Adult Social Care? This might be something the government, home care providers, training providers or regulators can do. |

**(b) Care home survey questions**

| Topic | Questions |
| --- | --- |
| Care home demographics | Care home name  Care home ICB location   a. North East London b. Kent and Medway c. Nottingham and Nottinghamshire d. North East and North Cumbria e. Lancashire and South Cumbria   Type of care provided |
| Participant demographics | Role  Duration in role   a. Less than one year b. 1-3 years c. 3-5 years d. 5+ years |
| Care home characteristics | Q1 Who owns the care home?   a. Private company b. Not for profit c. Council d. Charity e. Other   Q1 comment - if other, please specify  Q2 Is your care home independent or part of a chain?   a. Independent b. Chain   Q2 comment – what is the name of the care home?  Q3 How many care workers are employed at your care home?   a. Full time b. Part-time c. Agency/bank role   Q4 How many nurses are employed at your care home?   a. Full-time general nurses b. Part-time general nurses c. Full-time mental health nurses d. Part-time mental health nurses   Q5 How many activity coordinators (reported at FTE) are employed at your care home?  Q6 For each role, how many are employed at your care home?   a. Care worker b. Mental health nurse c. General nurse d. Activity coordinator e. Manager f. Other   Q6 comment – If other, please specify  Q7 Approximately, over the last 6 months…   a. How many new staff members (excluding agency) have joined the care home? b. How many staff members (excluding agency) have left the care home? |
| Care home occupants | Q8a What is the maximum occupancy of your care home?  Q8b How many places are currently unfilled?  Q8c Thinking about how clients fund their care, approximately how many (%) of your clients are…   a. Paying fully privately b. Part paying privately and part funded by the local authority c. Fully funded by the local authority   Q8d Approximately what proportion of residents are aged over 65?   a. Over 80% b. 60-80% c. 40-60% d. Under 40%   Q8e Approximately how many residents have a dementia diagnosis?   a. Almost all b. Most c. Some d. A few   Q8f Approximately what proportion of residents have some problems with memory or thinking that impact their ability to live independently?   a. Over 80% b. 60-80% c. 40-60% d. Under 40%   Q9 What are some of the common reasons why you were unable to accept people referred to you when you had space in the 6 months? |
| Training | Q10a What proportion of staff (%) receive dementia specific training?  Q10b Who provides this training, and how many hours are provided to staff?   a. In house b. An external supplier c. Other   Q10b comment – if other please specify  Q10c How is this training provided?   a. In person b. E-learning c. Video call d. Other   Q10c comment – if other please specify  Q11 Are there any training needs not covered by current dementia training that would like to cover?  12a How many activity groups are run per week?  12b What type of activities are offered?  12c Can residents take part in activities when the activity coordinator is not present?   a. Yes b. No c. Depends on the activity |
| Staff support | Q13 How are staff supported to deliver best practice care when residents show behaviours that challenge?   a. Use a specific model of care b. Regular multidisciplinary discussion groups c. Additional training specific to challenging behaviours d. Other   Q13 comment – if other please specify |
| Medication and medical input | Q14 For residents on anti-psychotic medication, is this regularly reviewed?   a. Yes b. No   Q14b Who is involved in this review?  Q15a How is medical input into the care home organised?   a. One GP who provides care to all residents b. Residents have their own GP   Q15b If one GP, do they visit on an ad-hoc basis as needed, or have visits?   a. Ad-hoc visits b. Regular visits   Q15c What professional visit the care home on a regular basis?   a. GP (ad-hoc or regular – how often) b. Geriatrician (ad-hoc or regular – how often) c. Psychiatrist (ad-hoc or regular – how often) d. Mental health nurse (ad-hoc or regular – how often) e. Physiotherapists (ad-hoc or regular – how often) f. Speech and language therapists (ad-hoc or regular – how often) g. Occupational therapists (ad-hoc or regular – how often) h. Other 1 (ad-hoc or regular – how often) i. Other 2 (ad-hoc or regular – how often)   Q15 comment – if other(s), please specify  Q16 Is there a regular MDT meeting?   a. Yes b. No   Q16b Who usually attend [MDT meeting]?   a. Care home manager b. GP or representative from local GP surgery c. Nurses d. Care workers e. Activity coordinators f. Specialist (e.g., dementia, end of life care) g. Other   Q16b comment – if other, please specify |
| Language challenges | Q17 Do you provide care to residents who do not speak English sufficiently enough to communicate their needs?   a. Yes b. No   Q17b How do you manage this?  Q18 Do you have access to an interpreter?   a. Yes, funded by the local authority b. Yes, paid privately c. No |
| Outcomes | Q19 What changes in how the care home operates in the last 5 years are you most proud of?  Q20 Are there any aspects of how your care home has operated in the last 5 years that concern you?  Q21 What changes, if any, do you anticipate in your care home over the next 5 years? |

Abbreviations

**Abbreviations:**

AO1: Assistance of One

ASC: Adult Social Care

CST: Cognitive Stimulation Therapy

CQC: Care Quality Commission

DN: District Nurse

GP: General Practitioner

HCA: Health Care Assistant

ICB: Integrated Care Board

LA: Local Authority

MDT: Multi Disciplinary Team Meeting

NHS: National Health Service

NICE: National Institute for Health and Care Excellence

OT: Occupational Therapist

PCN: Primary Care Network

PIR: Provider Information Return

UCP: Universal Care Plan
